# Disruption and recovery of notifiable infectious diseases after COVID-19 in Australia, 2015–2025

**DOI:** 10.64898/2026.02.13.26346301

**Authors:** Hayden Farquhar

## Abstract

**Background:** COVID-19 non-pharmaceutical interventions (NPIs) disrupted transmission of many infectious diseases worldwide. While disruption patterns are well-documented, systematic analysis of post-pandemic recovery trajectories across diverse pathogens remains limited. We examined disruption and recovery of 47 nationally notifiable diseases in Australia from 2015 to 2025.

**Methods:** We analysed NNDSS surveillance data for 47 diseases across six transmission modes, quantifying disruption using observed-to-expected (O/E) ratios against 2015–2019 baselines. We applied difference-in-differences (DiD) to estimate causal NPI effects, Kaplan-Meier survival analysis for time-to-recovery, and bootstrap 95% confidence intervals for cumulative immunity debt.

**Results:** During 2020–2021, 28 diseases decreased (median O/E 0.51), with border-sensitive and vaccine-preventable diseases most affected. DiD analysis estimated that border closures were associated with significantly greater suppression among import-dependent diseases (coefficient −0.50, 95% CI −0.90 to −0.10, p=0.016). By 2025, recovery was heterogeneous: 17 diseases exceeded baseline levels, 12 returned to expected levels, 15 remained below baseline (9 partially recovered, 6 in sustained suppression), and 3 had insufficient data for trajectory classification. Five diseases showed suppression-then-overshoot trajectories suggestive of immunity debt, though bootstrap 95% confidence intervals confirmed statistically significant cumulative excess for only one (rotavirus); for influenza, high baseline variability precluded statistical confirmation despite a large absolute overshoot.

**Conclusions:** Post-pandemic disease recovery in Australia is heterogeneous and incomplete. Fifteen of 47 diseases have not returned to baseline levels by 2025, while 17 exhibit overshoot. These findings argue for differentiated surveillance of still-suppressed diseases and targeted catch-up vaccination in pandemic birth cohorts.

**Article summary:** We analysed disruption and recovery of 47 nationally notifiable diseases in Australia from 2015 to 2025, finding that 15 diseases remain below pre-pandemic levels three years after NPI relaxation. Border closures caused disproportionate suppression of import-dependent diseases, and recovery trajectories varied by disease characteristics, with immunity debt statistically confirmed for only one of five candidate diseases.

## Introduction

Between 2020 and 2022, Australian governments deployed an array of non-pharmaceutical interventions (NPIs) — international border closures, domestic movement restrictions, physical distancing mandates, and enhanced hygiene practices — primarily targeting SARS-CoV-2 transmission. These measures simultaneously disrupted the epidemiology of numerous other infectious diseases through shared and collateral pathways.^1-3^

The collateral effects of NPIs have been documented across multiple countries.^2,7,8^ Three Australian studies addressed this directly: Haque et al. analysed 64 NNDSS diseases through 2022,^4^ Sohail et al. examined 68 diseases through 2022,^5^ and Rafferty et al. described trends during 2020–2021.^6^ These analyses established that NPIs reduced most respiratory and vector-borne diseases substantially, with reductions correlating with NPI intensity. What they did not address was what happened after NPIs were relaxed and borders reopened: which diseases return to pre-pandemic levels, which overshoot — consistent with “immunity debt,” the accumulation of susceptible individuals due to reduced pathogen exposure during the NPI period^14^ — and which remain suppressed, potentially reflecting lasting changes in behaviour, testing, or transmission dynamics?

We extended the analysis through 2025, capturing three full years of post-restriction data, and applied difference-in-differences (DiD), trajectory clustering, time-to-recovery survival analysis, and bootstrap uncertainty quantification — approaches not previously applied to the full Australian notifiable disease portfolio. Our aims were: first, to confirm disruption findings using a DiD framework providing stronger causal evidence than before-after comparisons; and second — the main contribution — to characterise recovery trajectories through 2025, including time-to-recovery, predictors of recovery speed, and whether cumulative immunity debt can be quantified with formal uncertainty bounds.

## Methods

### Data sources

We obtained aggregate notification data from Australia’s NNDSS via the Power BI dashboard (nndss.health.gov.au) for all nationally notifiable diseases: annual notification rates per 100,000 population by disease, state/territory, age group, and year (1991–2025), with ABS estimated resident population mid-year estimates as denominators. Data for 2025 were extracted in January 2026; all primary analyses were repeated excluding 2025 as a sensitivity analysis.^6^ Record-level datasets for influenza, meningococcal disease, and salmonellosis were obtained from cdc.gov.au, providing monthly resolution from 2008. State-level NPI stringency was derived from the Oxford COVID-19 Government Response Tracker subnational dataset.^9^

### Disease classification

Of 66 diseases with available data, 47 met inclusion criteria: complete notification data spanning baseline (2015–2019), COVID-acute (2020–2021), transition (2022), and post-COVID (2023–2025) periods. Nineteen diseases were excluded (Table S1). The 47 included diseases were classified into six mutually exclusive transmission modes (respiratory, n=14; enteric, n=12; vector-borne, n=6; zoonotic, n=5; sexually transmitted, n=5; blood-borne, n=5) and characterised as border-sensitive (n=9; requiring international importation for sustained transmission) or vaccine-preventable (n=12). The baseline period of 2015–2019 was selected to balance data completeness with temporal relevance (see supplementary material for rationale).

### Disruption quantification

For each disease, we computed observed-to-expected (O/E) ratios by dividing observed annual notification rates by the 2015–2019 baseline mean. We fitted individual interrupted time series (ITS) models for 44 diseases (excluding 3 with ultra-low counts): rate ∼ year + covid + time_since_covid, with Newey-West HAC standard errors^10,18^ (lag 1). With only 11 annual time points, these models are descriptive trend indicators. For three diseases with monthly data (influenza, meningococcal disease, salmonellosis), we fitted generalised least squares (GLS) models with AR(1) correlation and harmonic seasonal terms as the primary inferential ITS analyses.

We fitted a pooled mixed-effects model across all 47 diseases using z-scored rates, with random intercepts and random COVID effects by disease, testing whether disruption varied systematically by disease characteristics. Nested predictor regression models are reported in the supplementary material.

### Difference-in-differences

To estimate the causal effect of NPIs, we implemented a DiD design comparing border-sensitive diseases (n=9) with non-border-sensitive controls (n=38), exploiting the >98% reduction in international arrivals during 2020–2021 under the Biosecurity Act human biosecurity emergency declaration.^20^ Annual rates were z-scored within each disease, and we fitted linear mixed-effects models with random intercepts by disease. Pre-trend and placebo tests assessed the parallel trends assumption.^11^ A broader secondary specification (NPI-sensitive: respiratory or border-sensitive) is reported in the supplementary material.

### Time-to-recovery survival analysis

For the 28 diseases suppressed during COVID (O/E < 0.9), we defined recovery as the first year with O/E ≥ 0.9 and included survival analysis to visualise recovery timelines across diseases. We chose 0.9 rather than 1.0 to allow for normal inter-annual fluctuation; results were qualitatively similar with thresholds of 0.85 and 1.0. Diseases not recovered by 2025 were right-censored. We constructed Kaplan-Meier curves stratified by disruption severity group and compared distributions using log-rank tests. With only 16 events, Cox regression was exploratory.^13^

### Immunity debt quantification

For each disease-year, we computed cumulative deficit (expected minus observed) and cumulative excess (observed minus expected) relative to baseline. Net immunity debt was the difference over 2020–2025. We generated bootstrap 95% confidence intervals by resampling the 5 baseline years with replacement (999 iterations, percentile method). With only 5 values to resample, the bootstrap distribution is discrete (3,125 unique resamples) and these CIs should be interpreted as approximate; a parametric alternative using the t-distribution with 4 degrees of freedom would yield wider intervals.

### Sensitivity analyses

Five sensitivity analyses assessed robustness: (1) trend-adjusted baselines; (2) exclusion of 2025 data; (3) alternative COVID period boundary; (4) Poisson versus OLS comparison; (5) exclusion of high-variability diseases. Diseases were stratified by median COVID-acute O/E ratio, with k-medoids clustering^12^ as a secondary approach (see supplementary material for details).

All analyses were performed in R version 4.4.1.

## Results

### Disruption during COVID-19 (2020–2021)

Of 47 analysed diseases, 28 (60%) showed decreased notifications during the COVID-acute period, 10 (21%) increased, and 9 (19%) were unchanged (O/E between 0.9 and 1.1). The most disrupted were influenza (O/E 0.065, 93.5% reduction), dengue (O/E 0.076), and pertussis (O/E 0.115), all respiratory or border-sensitive. In individual ITS models, 26 of 44 diseases had significant COVID-period level changes (Newey-West HAC, p < 0.05); the pooled mixed-effects model provides the primary inferential result across diseases.

Disruption varied significantly by disease characteristics. Border-sensitive diseases had markedly lower O/E ratios than non-border-sensitive diseases (median 0.25 vs 0.86; Wilcoxon p=0.002). Vaccine-preventable diseases were more disrupted than non-VPD (median 0.29 vs 0.85; p<0.001). In the pooled mixed-effects model, the border sensitivity × COVID interaction was significant (coefficient −2.33, p=0.005); VPD status showed a borderline effect (p=0.07).

The border-sensitive DiD estimated significantly greater disruption among import-dependent diseases (coefficient −0.50, 95% CI −0.90 to −0.10, p=0.016; Table 2), with a non-significant pre-trend test supporting the parallel trends assumption. A broader NPI-sensitive specification yielded a similar coefficient (−0.44, p=0.022) but failed the pre-trend test (p=0.025), limiting its causal interpretability (supplementary material).

### Recovery trajectories (2022–2025)

Splitting diseases by median COVID-acute O/E ratio produced two groups with markedly different recoveries: the more-disrupted half (n=24, mean COVID O/E 0.35) had 55% still suppressed by 2025, while the less-disrupted half (n=23, mean O/E 0.94) had only 4% still suppressed. K-medoids clustering (k=2, silhouette width 0.235) produced a similar partition (Table 1).

**Table 1.** Characteristics of 47 nationally notifiable diseases by disruption severity group.

| Disease | TM | Bdr | VPD | N/yr | Rate | Cov OE | Post OE | Traj | Rec | Debt | Cl |
| --- | --- | --- | --- | --- | --- | --- | --- | --- | --- | --- | --- |
| Dengue | Vec | Y |  | 1510 | 6.20 | 0.08 | 1.08 | Returned | 2024 |  | 1 |
| Mumps | Resp |  | Y | 614 | 2.54 | 0.13 | 0.24 | Suppressed | — |  | 1 |
| Chikungunya | Vec | Y |  | 91 | 0.37 | 0.19 | 0.94 | Returned | 2025 |  | 1 |
| Rubella | Resp |  | Y | 15 | 0.06 | 0.25 | 0.22 | Suppressed | — |  | 1 |
| Flavivirus (unspec.) | Vec |  |  | 33 | 0.13 | 0.26 | 0.60 | Partial (slow) | — |  | 1 |
| Meningococcal | Resp |  | Y | 260 | 1.11 | 0.29 | 0.45 | Suppressed | — |  | 1 |
| Hepatitis E | Ent | Y |  | 44 | 0.18 | 0.39 | 0.56 | Partial (slow) | — |  | 1 |
| HUS | Ent |  |  | 16 | 0.07 | 0.59 | 0.83 | Partial | 2023 |  | 1 |
| Listeriosis | Ent |  |  | 70 | 0.29 | 0.59 | 0.93 | Returned | 2022 |  | 1 |
| Hep B (new) | BB |  | Y | 154 | 0.63 | 0.65 | 0.46 | Suppressed | — |  | 1 |
| Salmonellosis | Ent |  |  | 16017 | 65.40 | 0.68 | 0.68 | Partial (slow) | — |  | 1 |
| Hep C (unspec.) | BB |  |  | 9809 | 40.00 | 0.69 | 0.61 | Partial (slow) | — |  | 1 |
| Varicella (cpox) | Resp |  | Y | 3615 | 21.60 | 0.69 | 0.63 | Partial (slow) | — |  | 1 |
| Leprosy | Zoo | Y |  | 12 | 0.05 | 0.70 | 1.00 | Returned | 2021 |  | 1 |
| Hep B (unspec.) | BB |  |  | 5862 | 23.80 | 0.78 | 0.85 | Partial | — |  | 1 |
| Ross River | Vec |  |  | 5254 | 21.40 | 0.86 | 0.40 | Suppressed | — |  | 1 |
| Chlamydia | STI |  |  | 98915 | 402.20 | 0.87 | 0.95 | Returned | 2022 |  | 1 |
| H. influenzae b | Resp |  | Y | 18 | 0.07 | 0.95 | 0.45 | Suppressed | — |  | 1 |
| Tetanus | Zoo |  |  | 4 | 0.02 | 1.25 | 0.83 | Insuff. | — |  | 1 |
| Barmah Forest | Vec |  |  | 395 | 1.59 | 1.57 | 0.62 | Partial | — |  | 1 |
| Influenza | Resp |  | Y | 163005 | 658.80 | 0.07 | 2.20 | Major OS | 2022 | Y | 2 |
| Pertussis | Resp |  | Y | 15906 | 65.00 | 0.12 | 1.64 | Mod OS | 2024 | Y | 2 |
| Hepatitis A | Ent | Y | Y | 244 | 1.04 | 0.21 | 0.85 | Partial | — |  | 2 |
| Paratyphoid | Ent | Y |  | 84 | 0.34 | 0.25 | 1.21 | Mod OS | 2023 | Y | 2 |
| Malaria | Vec | Y |  | 338 | 1.40 | 0.30 | 1.19 | Returned | 2024 |  | 2 |
| Typhoid | Ent | Y |  | 148 | 0.60 | 0.33 | 1.53 | Mod OS | 2022 | Y | 2 |
| Rotavirus | Ent |  | Y | 4693 | 19.00 | 0.45 | 1.86 | Mod OS | 2022 | Y | 2 |
| Shigellosis | Ent | Y |  | 1972 | 7.80 | 0.51 | 1.41 | Mod OS | 2023 |  | 2 |
| Cryptosporidiosis | Ent |  |  | 3974 | 16.20 | 0.52 | 1.75 | Mod OS | 2024 |  | 2 |
| Pneumococcal | Resp |  | Y | 1869 | 7.40 | 0.61 | 1.26 | Mod OS | 2022 |  | 2 |
| Brucellosis | Zoo |  |  | 19 | 0.08 | 0.86 | 1.18 | Returned | 2023 |  | 2 |
| Q fever | Zoo |  |  | 545 | 2.20 | 0.91 | 1.21 | Mod OS | — |  | 2 |
| Gonorrhoea | STI |  |  | 27247 | 110.60 | 0.99 | 1.44 | Mod OS | — |  | 2 |
| Legionellosis | Resp |  |  | 401 | 2.00 | 1.00 | 1.50 | Mod OS | — |  | 2 |
| Tuberculosis | Resp |  |  | 1400 | 5.80 | 1.03 | 0.98 | Returned | — |  | 2 |
| Campylobacteriosis | Ent |  |  | 29120 | 136.40 | 1.04 | 1.11 | Returned | — |  | 2 |
| Syphilis (>=2yr) | STI |  |  | 2191 | 8.60 | 1.05 | 1.24 | Mod OS | — |  | 2 |
| Varicella (unspec.) | Resp |  |  | 13888 | 83.20 | 1.05 | 1.40 | Mod OS | 2021 |  | 2 |
| Hep C (new) | BB |  |  | 716 | 2.80 | 1.07 | 1.19 | Returned | — |  | 2 |
| Hepatitis D | BB |  |  | 67 | 0.27 | 1.15 | 1.58 | Mod OS | — |  | 2 |
| STEC | Ent |  |  | 440 | 1.72 | 1.17 | 2.14 | Major OS | — |  | 2 |
| Syphilis (<2yr) | STI |  |  | 4310 | 17.40 | 1.24 | 1.34 | Mod OS | — |  | 2 |
| Diphtheria | Resp |  | Y | 7 | 0.03 | 1.25 | 2.02 | Insuff. | — |  | 2 |
| Varicella (shingles) | Resp |  |  | 10696 | 63.40 | 1.30 | 0.97 | Returned | — |  | 2 |
| Leptospirosis | Zoo |  |  | 114 | 0.46 | 1.42 | 1.09 | Returned | 2021 |  | 2 |
| Ornithosis | Resp |  |  | 19 | 0.08 | 2.56 | 1.62 | Mod OS | — |  | 2 |
| Syphilis (cong.) | STI |  |  | 5 | 0.02 | 2.95 | 2.58 | Insuff. | — |  | 2 |

TM = transmission mode (Resp = respiratory, Ent = enteric, Vec = vector-borne, Zoo = zoonotic, STI = sexually transmitted, BB = blood-borne). Bdr = border-sensitive (Y = yes). VPD = vaccine-preventable (Y = yes). N/yr = mean annual notifications 2015–2019. Rate = mean annual rate per 100,000. Cov OE = mean observed/expected ratio during COVID-acute period (2020–2021). Post OE = mean observed/expected ratio during post-COVID period (2023–2025). Traj = recovery trajectory classification (see Methods). Rec = first year O/E returned to $\geq 0.9$ . Debt = immunity debt signal (Y = suppression followed by overshoot). Cl = disruption severity group from k-medoids partitioning (1 = heavily disrupted, 2 = mildly affected).

**Table 2.**
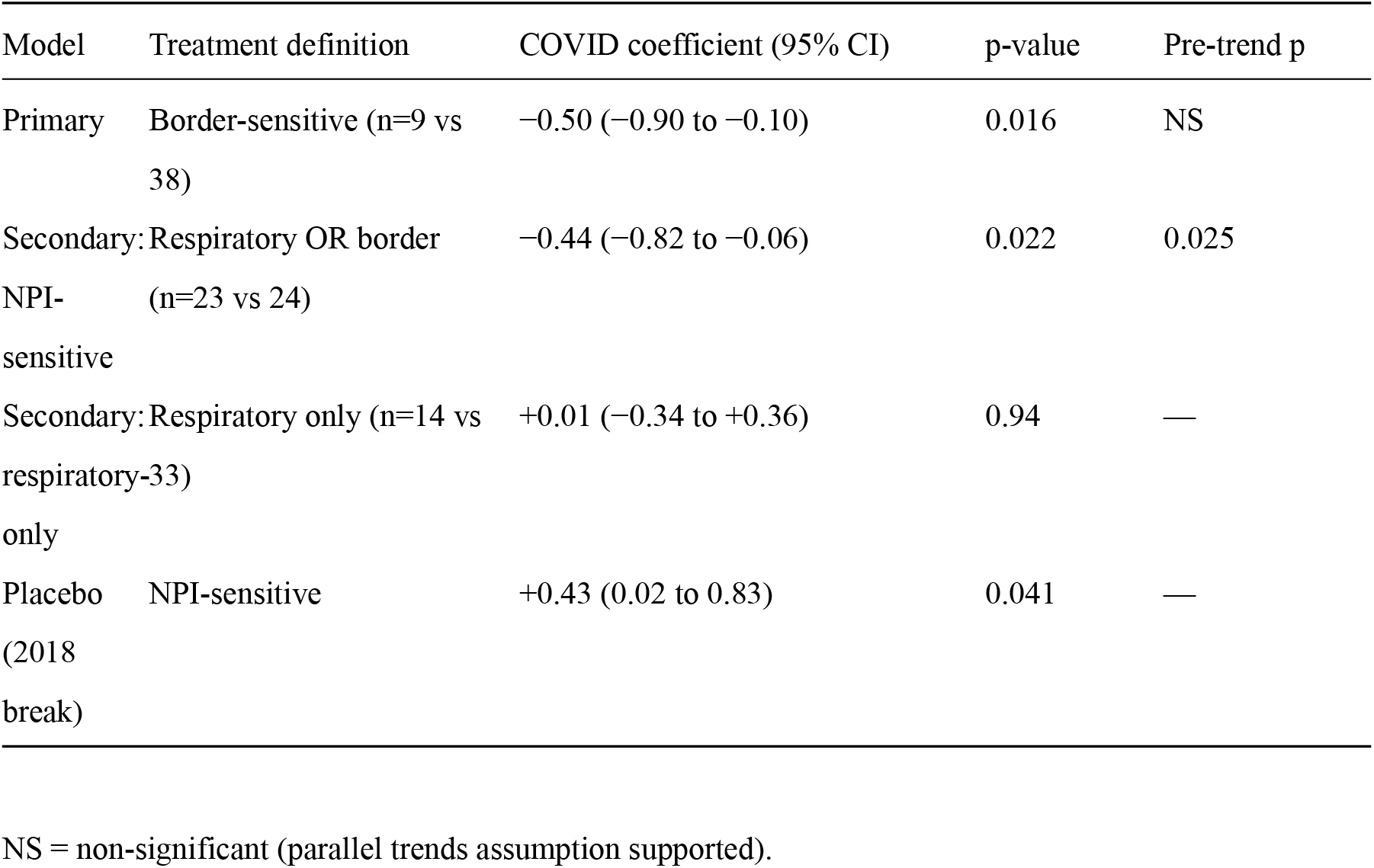
Difference-in-differences estimates.

Of 28 diseases suppressed during COVID (O/E < 0.9), 16 (57%) recovered by 2025 and 12 remained below baseline. Kaplan-Meier analysis showed a median time-to-recovery of 5 years from COVID onset, with recovery speed differing by disruption severity group (log-rank p=0.004) but not by border sensitivity (p=0.20) or VPD status (p=0.25). With only 16 recovery events, Cox regression was underpowered and no individual covariates reached significance. The significant log-rank result should be interpreted cautiously: more-disrupted diseases take longer to recover partly by definition, making this stratification partially tautological.

Using trajectory classification across all 47 diseases, 17 (36%) exhibited overshoot above baseline, 12 (26%) returned to expected levels, 9 (19%) partially recovered, and 6 (13%) remained in sustained suppression (3 diseases [6%] — tetanus, diphtheria, and congenital syphilis — had insufficient notification counts for trajectory classification but were retained in quantitative analyses). The 17 overshooting diseases include a mix of potential immunity debt (influenza, rotavirus), secular upward trends (gonorrhoea, syphilis), and other factors — overshoot alone does not indicate immunity debt. The most prominent overshoots were influenza (post-COVID O/E 2.20), Shiga toxin-producing *Escherichia coli* (STEC; O/E 2.14), and rotavirus (O/E 1.86).

Bootstrap analysis confirmed cumulative net excess (more post-COVID cases than deficit) for 24 diseases and net deficit for 23, with 31 of 47 (66%) confidence intervals excluding zero. The largest confirmed net excesses (CI excluding zero) were campylobacteriosis (+61,574 cases), gonococcal infection (+53,550), and varicella-unspecified (+29,423). The largest confirmed net deficits were salmonellosis (−27,761 cases), hepatitis C unspecified (−19,058), and varicella-chickenpox (−7,047). Influenza showed the largest absolute net excess (+435,226 cases) but with wide confidence intervals crossing zero (Table 3 footnote).

**Table 3.**
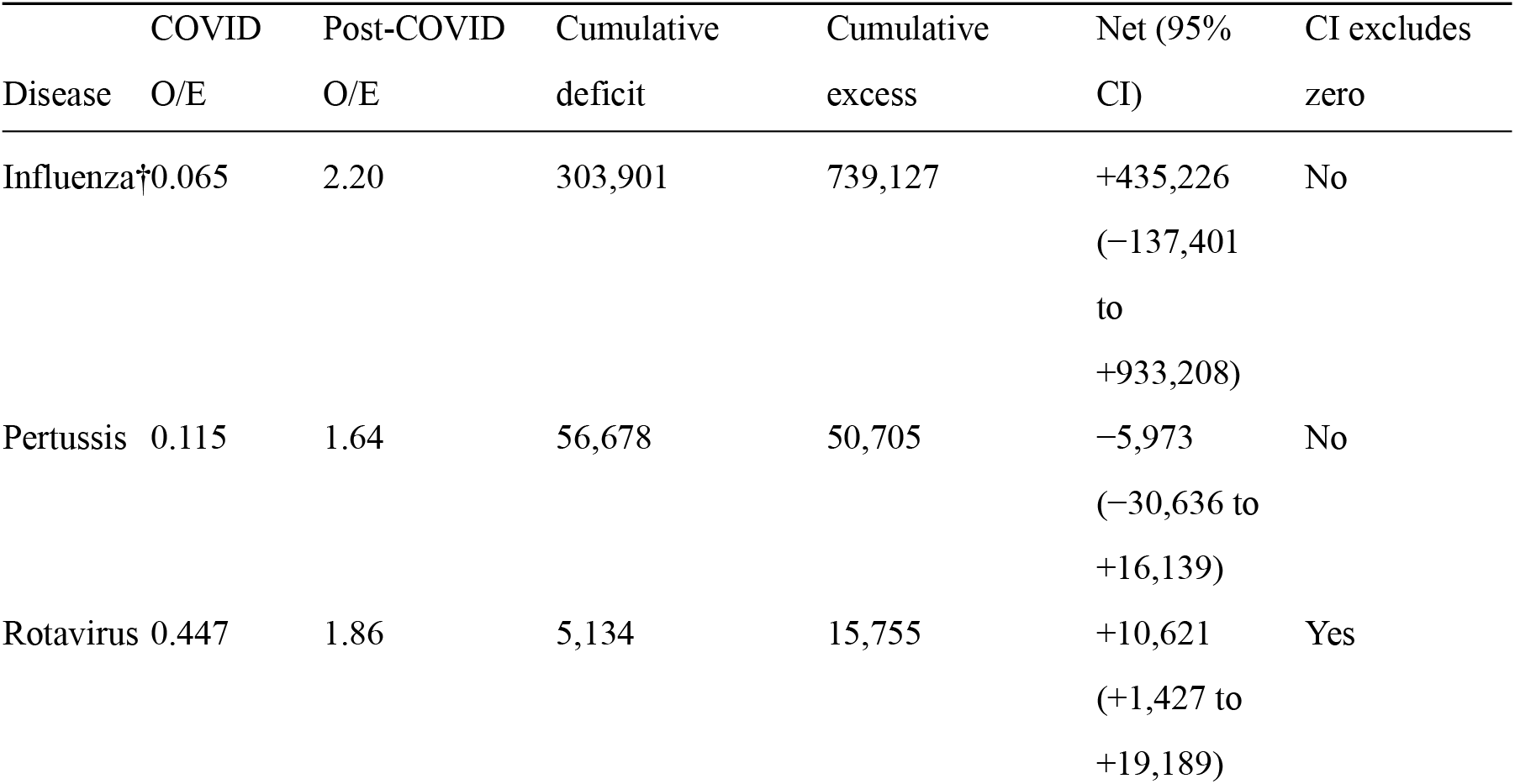
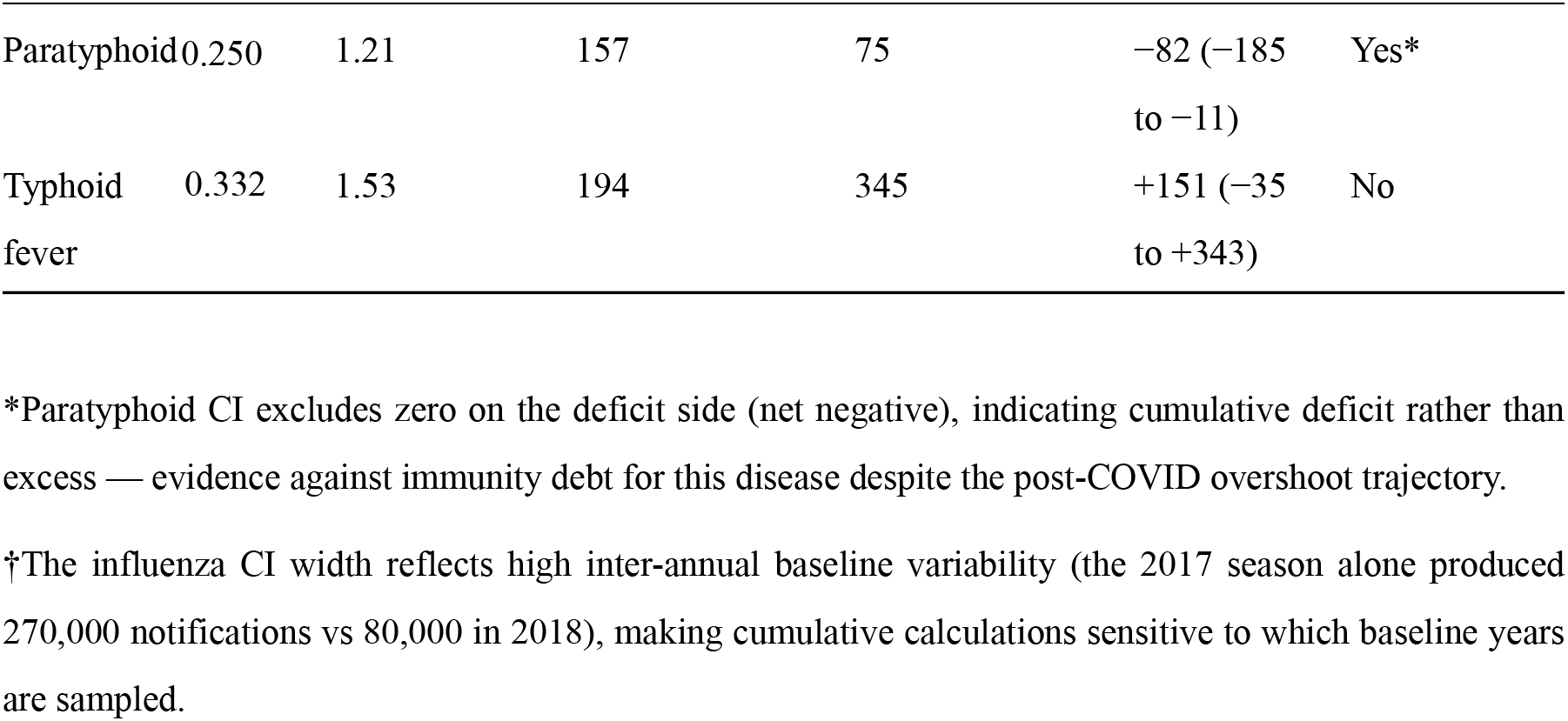
Immunity debt: diseases with suppression-overshoot trajectories.

Five diseases showed suppression-then-overshoot trajectories suggestive of immunity debt^14^ (Table 3). Of these, only rotavirus had a bootstrap CI excluding zero (+10,621 cases, 95% CI +1,427 to +19,189). For the remainder — including influenza, the most visually striking example — wide confidence intervals precluded statistical confirmation. Pertussis showed a near-balanced pattern: an 88% reduction during 2020– 2021 followed by 64% overshoot in 2024–2025, but the CI crossed zero. A recovery regression found that more-disrupted diseases had greater post-COVID O/E ratios (coefficient 0.41, p=0.01), though this is expected under regression to the mean — diseases with low COVID-period O/E ratios will tend to have higher subsequent ratios regardless of biological mechanism — and does not by itself support the immunity debt hypothesis.

### Sensitivity analyses

All five sensitivity checks confirmed the primary findings. Direction concordance ranged from 91% (Poisson vs OLS comparison) to 98% (2025 exclusion). Trend-adjusted baselines shifted O/E ratios for gonorrhoea and syphilis by 15–20% but did not alter disruption rankings. Full results are in the supplementary material.

## Discussion

Recovery from COVID-era disruption is heterogeneous and incomplete. Of the 28 diseases suppressed during the COVID-acute period, 16 (57%) had recovered to O/E ≥ 0.9 by 2025, while 12 (43%) remain below prepandemic rates three years after NPI relaxation.

### Recovery patterns and immunity debt

The split between heavily disrupted diseases with slow recovery and mildly affected diseases with rapid rebound is broadly consistent with immunity debt theory,^14^ but the statistical evidence is weaker than visual trajectories suggest. Of five diseases with classic suppression-then-overshoot patterns, only rotavirus had a bootstrap CI for cumulative excess that excluded zero. Influenza — the most dramatic example, with notifications at 2.2 times baseline in 2023–2024 — had such high pre-pandemic variability that the cumulative calculation cannot distinguish a genuine surplus from normal fluctuation. Pertussis came close: the 88% suppression followed by 64% overshoot is hard to explain without accumulated susceptibility, but the CI crossed zero.

This matters because “immunity debt” has become a widely invoked post-pandemic explanation, and our data show it is easier to see in rate trajectories than to confirm in cumulative case counts. The 31 diseases with CIs excluding zero are mostly those with low baseline variance, where even modest deviations are detectable — not necessarily where debt is most epidemiologically meaningful.

What the data support more clearly is the practical consequence: pertussis vaccination coverage dropped during the pandemic,^16,19^ and the affected birth cohorts (2020–2022) are now driving the overshoot. Whether one calls this “immunity debt” or “catch-up transmission in under-vaccinated cohorts” is partly semantic, but the implication for catch-up vaccination is the same.^14,15^

### Sustained suppression

Six diseases remained below 80% of baseline by 2025. The respiratory group — meningococcal disease and mumps — mirrors international trends; Brueggemann et al. reported sustained reductions in invasive meningococcal and pneumococcal disease across 26 countries.^17^ Whether reduced crowding and improved hygiene explain a 40–60% sustained reduction in Australia remains untested. An alternative is that reduced circulation disrupted natural immune boosting, creating a susceptibility reservoir that could eventually produce a rebound. Salmonellosis (sustained 50% reduction) more plausibly reflects durable changes in food handling practices. Ross River virus reductions likely reflect vector ecology and rainfall patterns rather than human behaviour.

Not all suppression is equal. If meningococcal carriage remains lower, that is a benefit. But if pertussis or measles circulation stays low because natural boosting has declined, the gap will eventually close through an outbreak rather than gradual return.^14,15^ Eight of 15 still-suppressed diseases show year-on-year O/E increases since 2022; seven show no upward trend — these warrant closest surveillance attention.

### Causal inference

The border-sensitive DiD provides the strongest causal evidence in this study, exploiting a clean mechanistic pathway — near-complete cessation of international arrivals — and passing pre-trend validation. The broader NPI-sensitive specification failed its pre-trend test, revealing that this grouping mixes diseases with divergent pre-pandemic trajectories (supplementary material). This heterogeneity means that prior studies using broad “respiratory disease” categories as a treatment group may be subject to similar violations.

### Limitations

The most important limitation is statistical power. Annual ITS models have only 11 time points and should be read as descriptive trends; the monthly models for influenza, meningococcal disease, and salmonellosis carry substantially more weight. The survival analysis had 16 events across 28 diseases, limiting Cox regression to exploratory univariate analysis. Surveillance artefacts are also a concern: changes in testing behaviour, healthcare-seeking, and laboratory capacity during the pandemic could confound observed trends, particularly for diseases diagnosed through routine screening (chlamydia, hepatitis C). The 2025 data may be affected by late notifications, though the sensitivity analysis excluding 2025 produced no qualitative changes. All analyses are ecological: we can describe disease-level patterns but cannot directly observe individual-level immunological mechanisms.

### Policy implications

First, the 15 diseases still below baseline warrant differentiated surveillance responses. Diseases on flat or declining trajectories (meningococcal disease, mumps, rubella) raise questions about whether circulation has permanently shifted, creating vulnerability to future outbreaks. Enhanced seroprevalence monitoring would help distinguish genuine elimination progress from dangerous immunity gaps.

Second, the disproportionate rebound in young children (Figure S8) — influenza in 0–4 year-olds reached 2.4 times baseline versus 1.1 in adults over 65 — argues for targeted catch-up strategies.^16^ Jurisdictions should audit coverage rates in 2020–2022 birth cohorts and consider supplementary vaccination campaigns where gaps are identified.

Third, the border-sensitive DiD finding (coefficient −0.50) has relevance for pandemic preparedness. If border closures are deployed again, planners should anticipate subsequent rebound in import-dependent diseases, including pre-positioned sentinel surveillance at points of entry.

Finally, diseases showing sustained overshoot (influenza, pertussis, STEC) are placing ongoing pressure on clinical and laboratory services. Surveillance planning should account for elevated notification rates persisting for several more years.

## Conclusions

Three years after Australia relaxed its pandemic restrictions, the infectious disease landscape has not returned to its pre-2020 state — and whether the current patterns represent permanent shifts or ongoing transients remains to be determined. Fifteen of 47 diseases remain below pre-pandemic rates, 17 now exceed baseline, and the statistical evidence for cumulative immunity debt is weaker than the visually striking overshoot trajectories imply. Whether these fifteen still-suppressed diseases represent a durable shift or a slow-burning accumulation of susceptibility will only become clear with continued monitoring. The most actionable signal is the disproportionate rebound in young children — the 2020–2022 birth cohorts who missed years of natural immune priming — which argues for targeted catch-up vaccination rather than waiting for routine schedules to close the gap.

## Data availability

All data used in this analysis are publicly available from the Australian National Notifiable Diseases Surveillance System (nndss.health.gov.au) and the Oxford COVID-19 Government Response Tracker (github.com/OxCGRT). Analysis code and processed data are available at https://github.com/hayden-farquhar/NNDSS-COVID-disruption.

## Funding

This research received no specific grant from any funding agency in the public, commercial, or not-for-profit sectors.

## Conflicts of interest

The author declares no conflicts of interest.

## Ethics statement

Ethics approval was not required as all data used are publicly available, de-identified, and aggregated at the disease–jurisdiction–year level. No individual-level data were accessed.

## AI use disclosure

AI-assisted tools (Claude, Anthropic) were used for code development, statistical verification, and manuscript drafting assistance. All analyses were designed, validated, and interpreted by the author. The author takes full responsibility for the content.

## Acknowledgements

The author acknowledges the Australian Government Department of Health and Aged Care for maintaining the National Notifiable Diseases Surveillance System (NNDSS) and making aggregate notification data publicly available through the NNDSS Power BI dashboard. Record-level datasets for influenza, meningococcal disease, and salmonellosis were obtained from the Australian Government Department of Health via the cdc.gov.au data portal. State and territory NPI stringency data were derived from the Oxford COVID-19 Government Response Tracker (OxCGRT), maintained by the Blavatnik School of Government, University of Oxford. Population denominators were sourced from the Australian Bureau of Statistics (ABS) estimated resident population (ERP) series.

---

**Figure 1.**
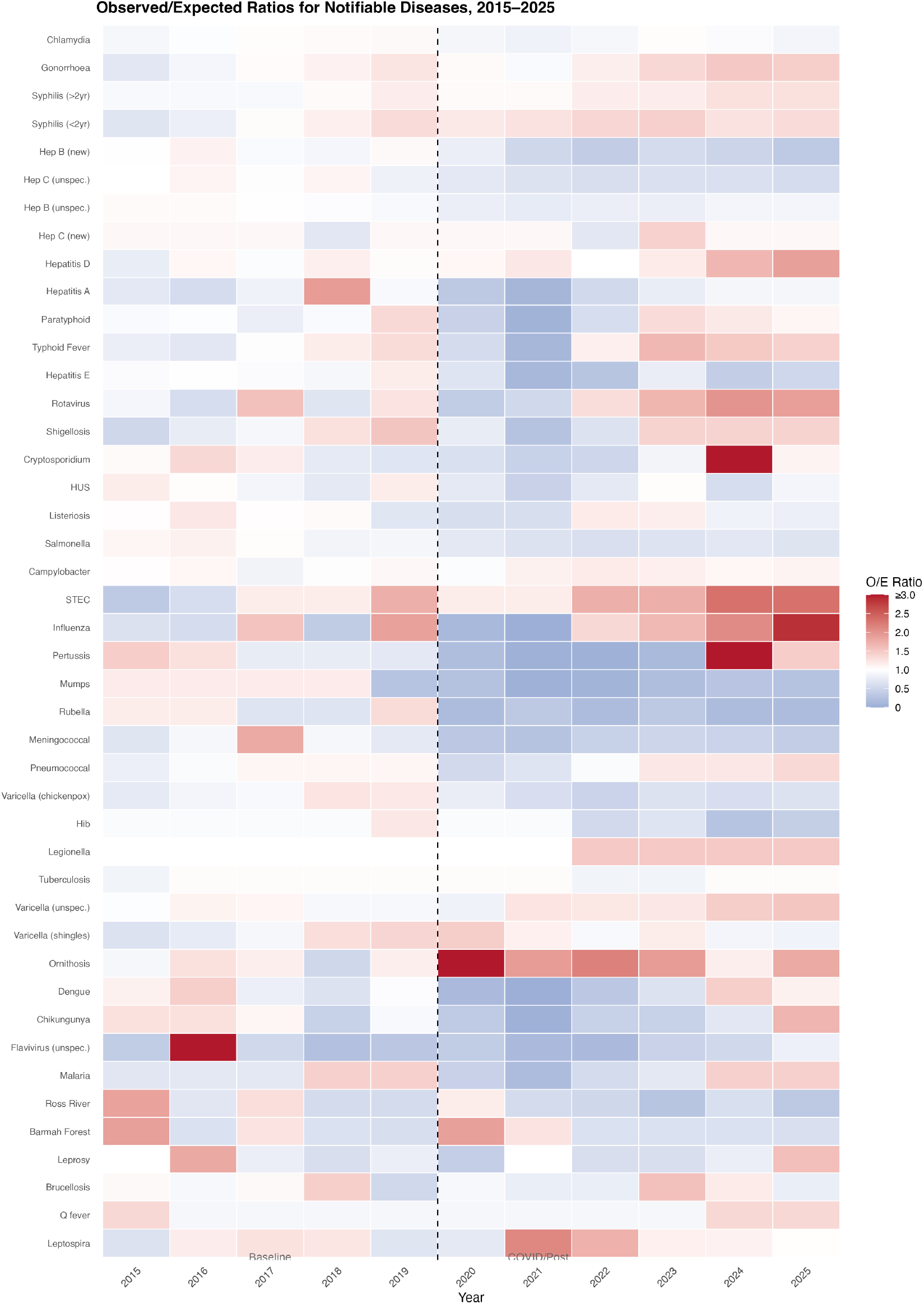
Heatmap of observed-to-expected (O/E) ratios for 47 nationally notifiable diseases in Australia, 2015–2025. Diseases are grouped by transmission mode and ordered by COVID-acute disruption magnitude. Blue indicates suppression (O/E < 1), red indicates overshoot (O/E > 1), white indicates baseline levels.

**Figure 2.**
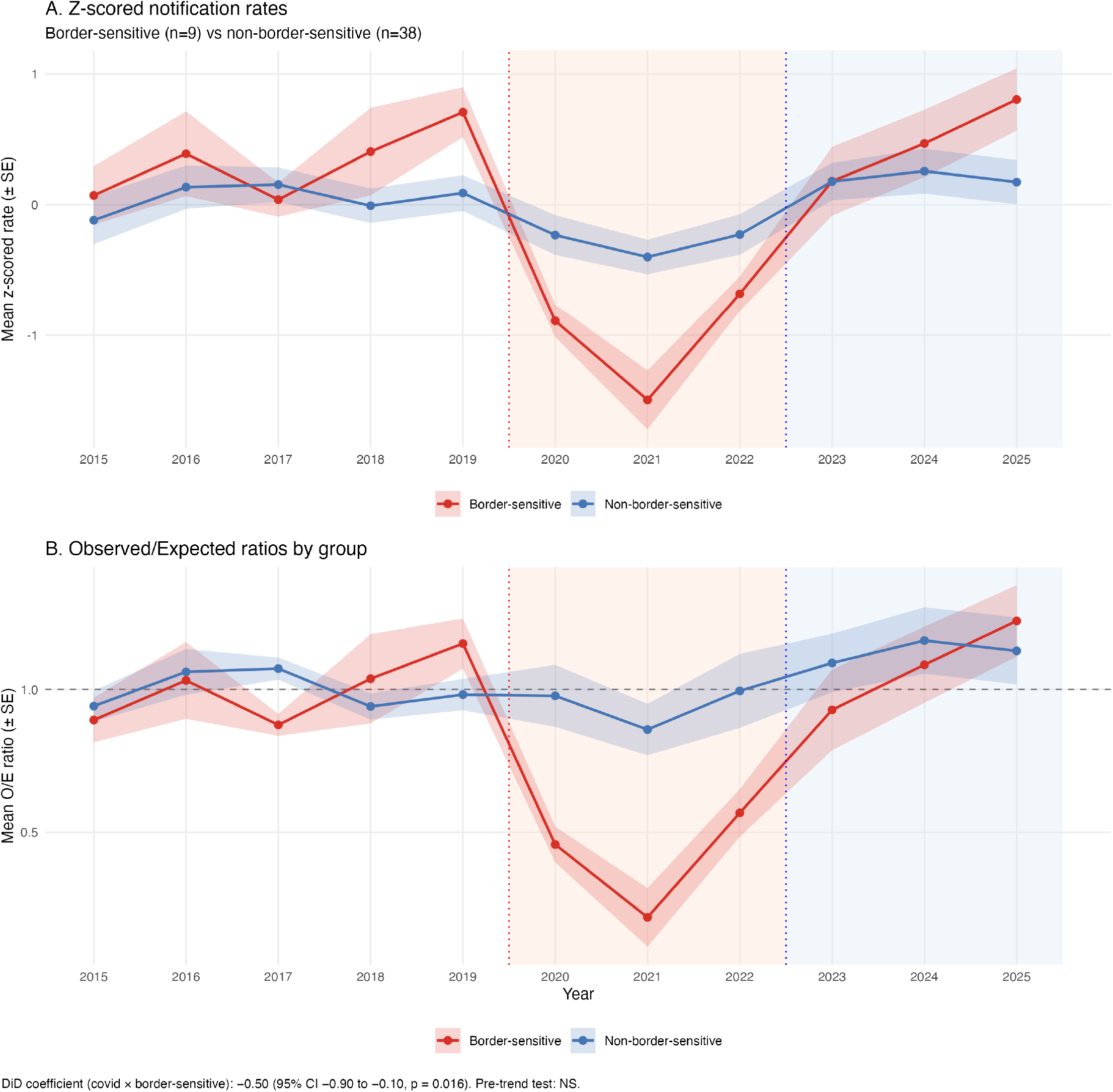
Difference-in-differences analysis (border-sensitive) Difference-in-differences analysis comparing border-sensitive diseases (n=9) with non-border-sensitive controls (n=38). Panel A shows mean z-scored notification rates; Panel B shows mean O/E ratios. The border-sensitive specification (coefficient −0.50, 95% CI −0.90 to −0.10, p=0.016) passed the pre-trend test.

**Figure 3.**
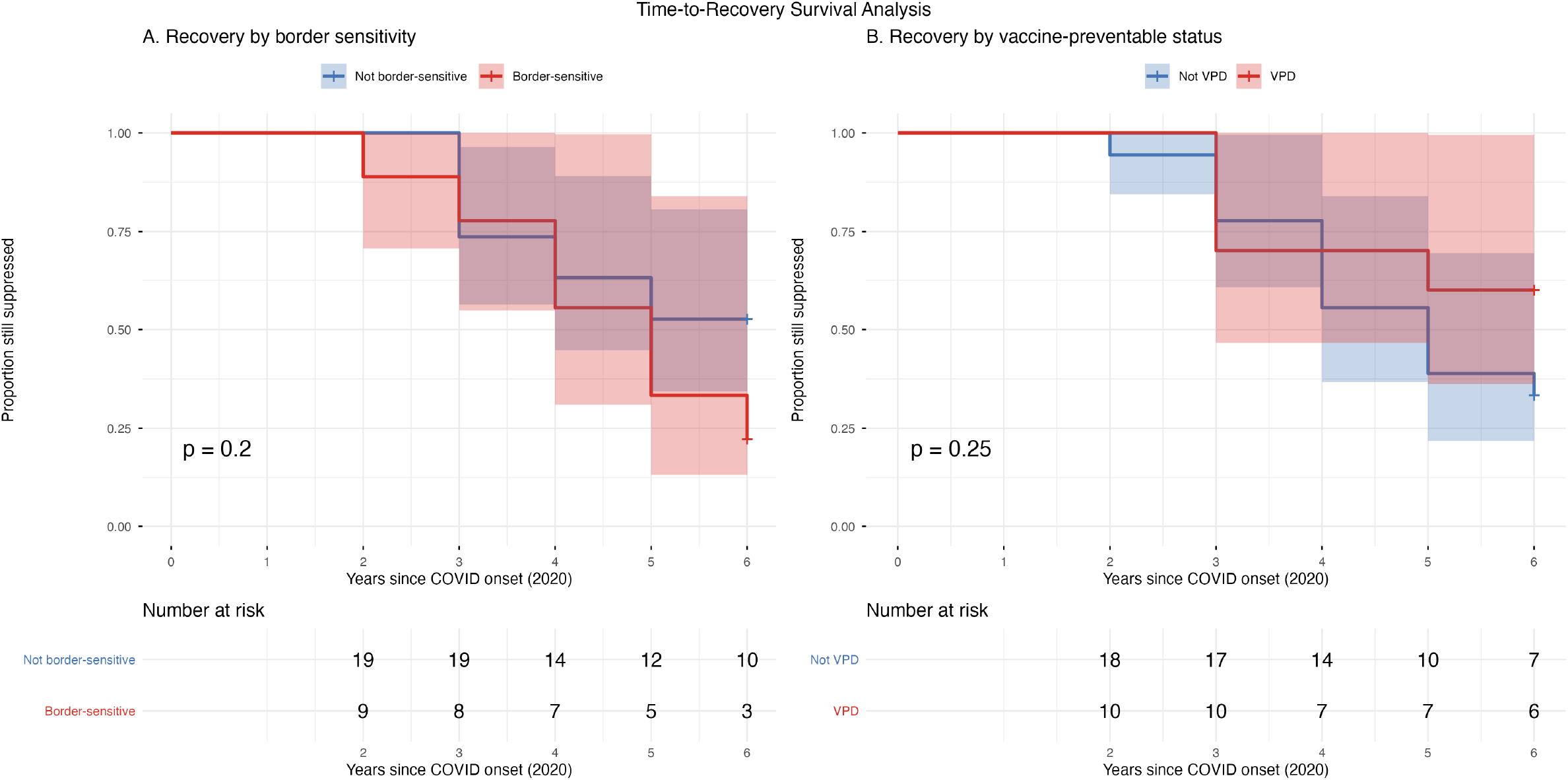
Kaplan-Meier curves for time-to-recovery among 28 diseases suppressed during COVID-19 (O/E< 0.9), stratified by disruption severity group. Recovery defined as first year with O/E ≥ 0.9; diseases not recovered by 2025 are right-censored. Log-rank p=0.004.

**Figure 4.**
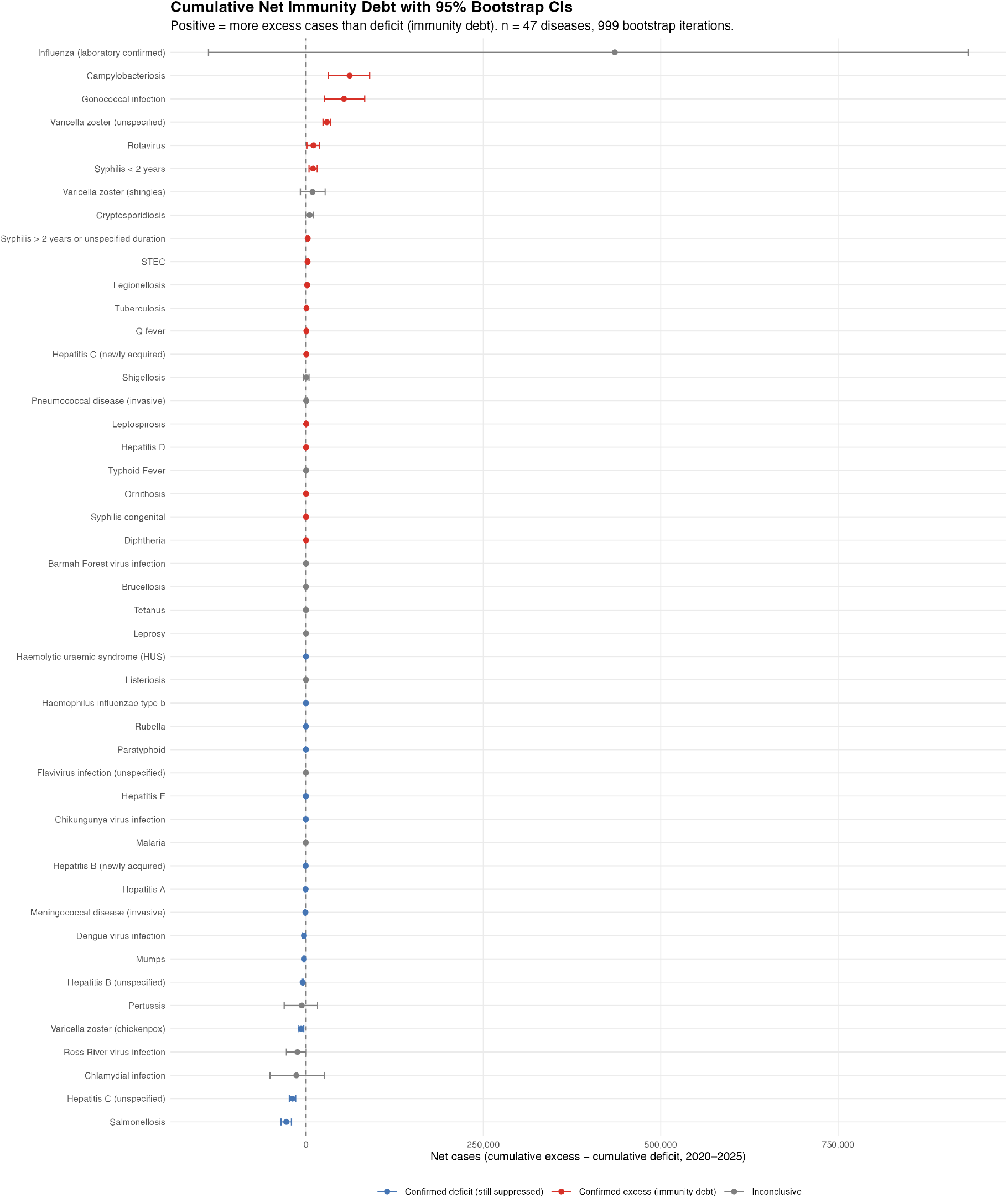
Immunity debt forest plot with bootstrap 95% Cis. Forest plot of cumulative net immunity debt (excess minus deficit cases, 2020–2025) with bootstrap 95% confidence intervals for 47 diseases. Filled circles indicate CIs excluding zero. Of 47 diseases, 31 have statistically significant cumulative imbalances.

## Notes

### Competing Interest Statement

The authors have declared no competing interest.

### Author Declarations

Ethics approval was not required as all data used are publicly available, de-identified, and aggregated at the disease-jurisdiction-year level. No individual-level data were accessed.

