## Supplementary for "Disruption and recovery of notifiable infectious diseases after COVID-19 in Australia, 2015–2025"

Hayden Farquhar, MBBS MPHTM

---

### Supplementary Methods

#### Baseline period selection

The baseline was set at 2015–2019 rather than extending further back for three reasons: (i) several diseases were added to or removed from the notification schedule between 2010 and 2014, so a longer window would reduce the number of diseases with complete data; (ii) secular trends in surveillance practices — including the transition to electronic laboratory reporting, advocated by Muscatello et al. in 2008[S1] and progressively implemented across jurisdictions during 2010–2013 — make earlier years less comparable; and (iii) a five-year window provides sufficient observations for stable mean estimates while remaining recent enough to reflect contemporary transmission dynamics. Trend-adjusted baselines for the 13 diseases with significant secular trends within 2015–2019 were assessed as a sensitivity analysis.

#### Predictor analysis

Nested regression models predicted COVID-period O/E ratios from disease characteristics. Five models were compared:

- **M1:** Transmission mode only
- **M2:** M1 + border sensitivity
- **M3:** M2 + vaccine-preventable (VPD) status
- **M4:** M3 + log baseline rate
- **M5 (parsimonious):** Border sensitivity + VPD status + log baseline rate (omitting transmission mode)

Models were compared by AIC, BIC, and adjusted $R^2$. Variance inflation factors (VIF) were computed for all predictors; all values were below 1.3, indicating no multicollinearity. The BIC-preferred model was M5

($R^2$=0.41), while M4 had the highest $R^2$ (0.54) but was penalised for overfitting by BIC. Border sensitivity and VPD status were the strongest predictors across all specifications.

A recovery regression tested whether disruption magnitude predicted post-COVID O/E ratios. More-disrupted diseases had greater post-COVID O/E ratios (coefficient 0.41, p=0.01), consistent with immunity debt but also expected under regression to the mean. Distinguishing these mechanisms would require simulation or Bland-Altman analysis.

**Disruption stratification: k-medoids clustering**

As a secondary approach to the primary median-split stratification, k-medoids clustering (PAM algorithm) was applied to COVID-acute O/E ratios. Silhouette widths were computed for k=2 through k=6; k=2 was optimal (silhouette width 0.235). The resulting partition — Cluster 1 (n=20, heavily disrupted, mean O/E 0.35) and Cluster 2 (n=27, mildly affected, mean O/E 0.94) — was similar to the median-split but not identical. The low silhouette width confirms a continuous spectrum of disruption rather than discrete categories.

**Secondary difference-in-differences specification**

In addition to the primary border-sensitive DiD, we tested a broader treatment definition classifying diseases as "NPI-sensitive" if they were either respiratory-transmitted or border-sensitive (n=23 treatment, n=24 control). This specification yielded a coefficient of −0.44 (95% CI −0.82 to −0.06, p=0.022), but failed the pre-trend test (p=0.025), indicating that these disease groups were already on divergent trajectories before the pandemic. A placebo test using a false 2018 break point was also significant (p=0.041), further undermining causal interpretation. We additionally tested a respiratory-only specification (n=14 vs 33), which was not significant (+0.01, p=0.94). We report the NPI-sensitive and respiratory-only results as secondary specifications that do not support causal inference.

The pre-trend failure is itself informative: it reveals that this broader grouping mixes diseases that declined sharply during COVID (influenza, pertussis) with others that increased (legionellosis, ornithosis), creating divergent pre-trends. Prior studies using broad "respiratory disease" categories as a treatment group may be subject to similar violations, even when untested.

**Immunity debt bootstrap detail**

For each disease-year, cumulative deficit (expected minus observed, when observed < expected) and cumulative excess (observed minus expected, when observed > expected) were computed over 2020–2025. Net immunity debt was the difference between cumulative excess and cumulative deficit. Bootstrap 95% confidence intervals were generated by resampling the 5 baseline years (2015–2019) with replacement (999 iterations), recomputing the baseline mean for each resample, and recalculating all debt metrics. The percentile method was used for CI construction. With only 5 values to resample, the bootstrap distribution is discrete ($5^5$ = 3,125 unique resamples) and these CIs should be interpreted as approximate. A parametric alternative using the t-distribution with 4 degrees of freedom would yield wider intervals; the bootstrap CIs may therefore be slightly anti-conservative.

**Sensitivity analyses**

Five sensitivity analyses assessed the robustness of primary findings:

1. **Trend-adjusted baselines:** For 13 diseases with statistically significant secular trends within 2015–2019 (linear regression $p < 0.05$), O/E ratios were recomputed using the trend-projected expected value rather than the five-year mean. This shifted O/E ratios for gonorrhoea and syphilis by 15–20% but did not alter disruption rankings or trajectory classifications.

2. **Exclusion of 2025 data:** Because 2025 data were extracted in January 2026 and may be affected by late notifications, all primary analyses were repeated using 2015–2024 data only. Direction concordance was 98%, and no qualitative changes to DiD or ITS coefficients were observed.

3. **Alternative COVID period boundary:** The primary analysis defined COVID-acute as 2020–2021; this sensitivity used 2020–2022 (including the transition year). Results were qualitatively unchanged; the broader definition slightly diluted effect sizes for diseases that began recovering in 2022.

4. **Poisson versus OLS comparison:** For 44 diseases, Poisson regression was fitted alongside the primary OLS ITS. Direction concordance was 91%; discordant cases were all diseases with O/E ratios near 1.0 where both models produced non-significant results.

5. **Exclusion of high-variability diseases:** Diseases with coefficient of variation (CV) > 0.5 during the baseline period were excluded (n=8). Direction concordance was 95% among the remaining 39 diseases.

Across all five checks, primary findings were robust: direction concordance ranged from 91% to 98%, and no sensitivity analysis changed the interpretation of the DiD, survival, or bootstrap results.

---

#### Table S1. Diseases excluded from analysis

Nineteen of 66 nationally notifiable diseases were excluded from the primary analysis. Exclusion reasons are summarised below.

| Disease | Transmission mode | Exclusion reason |
|---|---|---|
| Measles | Respiratory | Missing COVID-period data |
| Rubella (congenital) | Respiratory | Insufficient baseline (<5 years) |
| RSV | Respiratory | Notifiable from 2020 only |
| iGAS | Respiratory | Notifiable from 2021 only |
| Cholera | Enteric | Insufficient baseline (<5 years) |
| Botulism | Enteric | Insufficient baseline (<5 years) |
| *Vibrio parahaemolyticus* | Enteric | Notifiable from 2025 only |
| Donovanosis | STI | Insufficient baseline (last notified 2014) |
| Murray Valley encephalitis | Vector-borne | Insufficient baseline (<5 years) |
| Japanese encephalitis | Vector-borne | Insufficient baseline (<5 years) |
| West Nile/Kunjin | Vector-borne | Insufficient baseline (<5 years) |
| Hepatitis (NEC) | Blood-borne | Category discontinued |
| Anthrax | Zoonotic | Insufficient baseline (last notified 2010) |
| Australian bat lyssavirus | Zoonotic | No confirmed notifications |
| Tularaemia | Zoonotic | Insufficient baseline (<5 years) |
| Avian influenza in humans | Zoonotic | No confirmed notifications |

| Disease | Transmission mode | Exclusion reason |
|---|---|---|
| Poliovirus infection | Other | Insufficient baseline (last notified 2007) |
| COVID-19 | Other | Pandemic pathogen (the exposure of interest) |
| Mpox | Other | Emerged 2022 (no pre-COVID baseline) |

---

### Supplementary Figures

**Figure S1.** Annual notification time series for 44 notifiable diseases with sufficient data, 2015–2025 (3 diseases with ultra-low counts excluded from time series display). Each panel shows observed annual notification rates per 100,000 population with the 2015–2019 baseline mean (horizontal dashed line) and 95% prediction interval (shaded band). Diseases are grouped by transmission mode.

**Figure S2.** Forest plot of COVID-acute O/E ratios by disease. Horizontal bars show the observed-to-expected ratio for each disease during 2020–2021, ordered by magnitude. The vertical dashed line at O/E = 1.0 indicates no change from baseline. Diseases to the left were suppressed; those to the right increased. Border-sensitive diseases (filled markers) cluster at the low end.

**Figure S3.** Monthly time series with ARIMA baseline forecasts for influenza, meningococcal disease, and salmonellosis. Black lines show observed monthly notifications; blue dashed lines show ARIMA-forecasted expected counts; shading indicates 95% prediction intervals. Vertical lines mark COVID onset (March 2020) and border reopening (March 2022).

**Figure S4.** State-level comparison of disruption magnitude. Boxplots show the distribution of COVID-acute O/E ratios across diseases within each state/territory. Victoria was the most disrupted (median O/E 0.56), Western Australia the least (0.87), and the Northern Territory was an outlier (1.18).

**Figure S5.** Recovery trajectory classifications for all 47 diseases. Each panel shows the annual O/E ratio trajectory from 2015 to 2025 with diseases classified into five categories: major overshoot, moderate overshoot, returned, partial recovery, and sustained suppression. Background shading indicates the COVID-acute (2020–2021) and transition (2022) periods.

**Figure S6.** Monthly interrupted time series for influenza, meningococcal disease, and salmonellosis. Each panel shows the fitted GLS AR(1) model with harmonic seasonal terms, with the step and slope change at COVID onset. Influenza had a near-unit-root AR(1) parameter (phi=0.917), which absorbed the level shift and reduced the statistical significance of the COVID coefficient.

**Figure S7.** State-level NPI stringency versus median disease O/E ratio. Mean stringency index (from OxCGRT subnational data) is plotted against median COVID-acute O/E ratio for eight states/territories. The Spearman rank correlation was negative (rho = −0.38) but non-significant (p=0.35, n=8), reflecting limited statistical power with only eight observations.

**Figure S8.** Age-specific disruption and recovery for influenza and pneumococcal disease, 2015–2024. Children aged 0–4 show substantially greater post-pandemic overshoot than older age groups (influenza: 2.4× baseline in 0–4 year-olds vs 1.1× in adults over 65). Influenza 2025 age-specific data excluded due to incomplete record-level extract.

**Figure S9.** Difference-in-differences analysis comparing NPI-sensitive diseases (n=23) with controls (n=24). Panel A shows mean z-scored rates; Panel B shows mean O/E ratios. This broader specification yielded a coefficient of −0.44 (p=0.022) but failed the pre-trend test (p=0.025), limiting causal interpretability.

**Figure S10.** Disruption severity group profiles from k-medoids partitioning (k=2, silhouette width 0.235). Cluster 1 (n=20): heavily disrupted, mean COVID-acute O/E 0.35. Cluster 2 (n=27): mildly affected or overshoot, mean COVID-acute O/E 0.94.

**Figure S11.** Generalised additive model (GAM) recovery curves for influenza, meningococcal disease, and salmonellosis. The GAM improved fit for meningococcal disease (RMSE 0.333 vs ITS 0.368) but not for influenza or salmonellosis, indicating selective utility of non-linear modelling for recovery characterisation.

---

### Supplementary Figures (embedded)

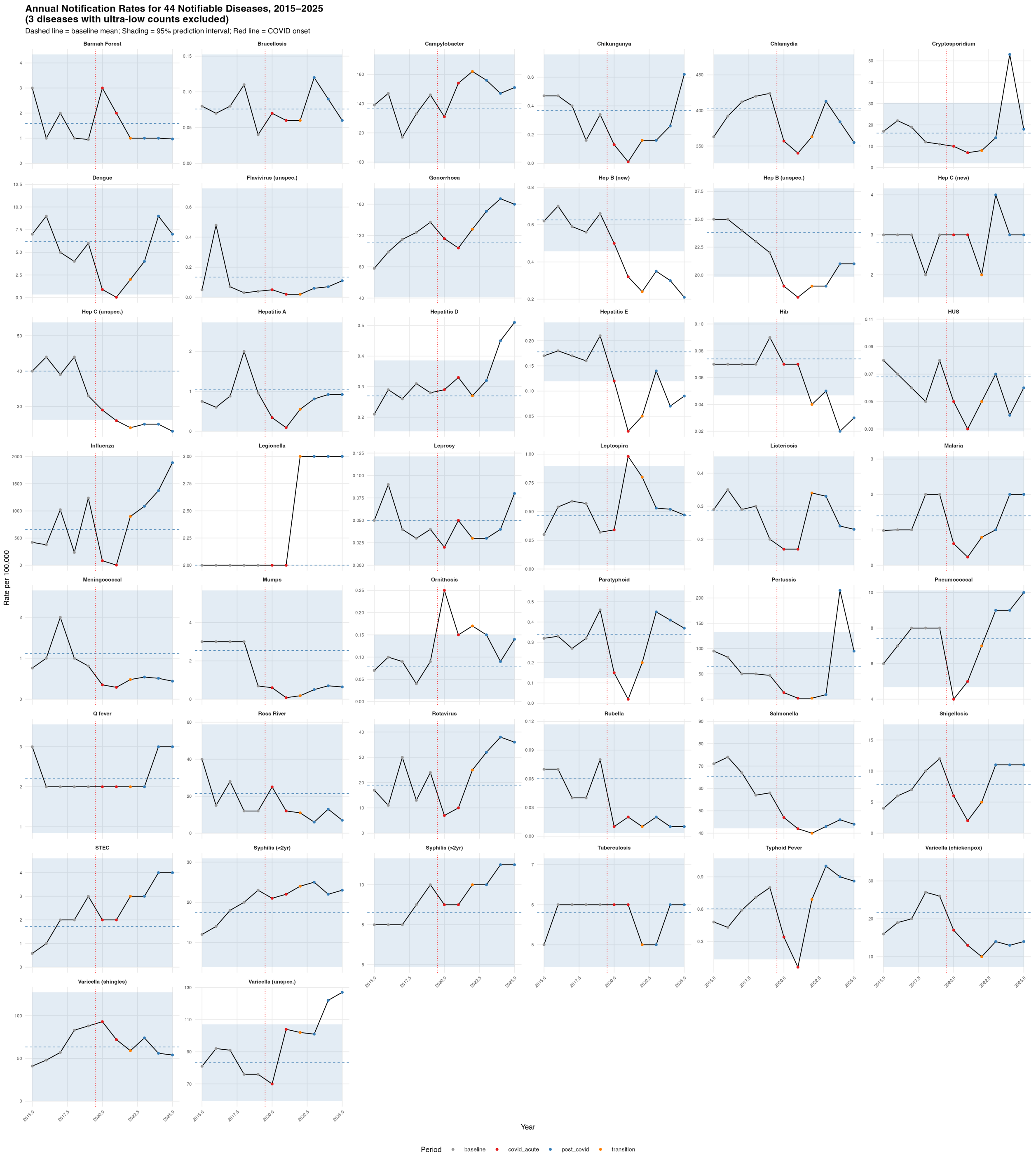


**Figure S1:** Figure S1. Annual time series for all 47 diseases

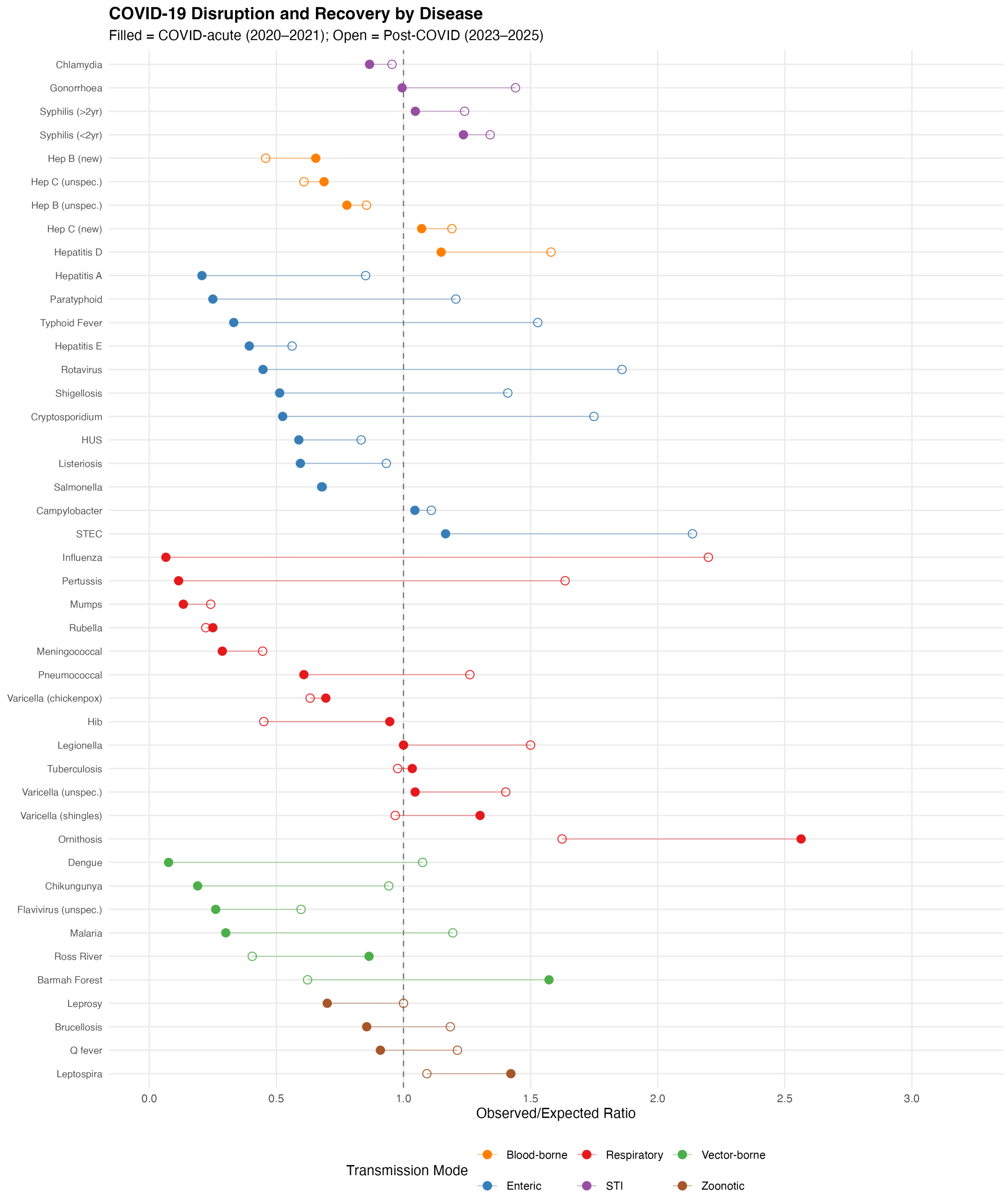


**Figure S2:** Figure S2. Forest plot of O/E ratios by disease

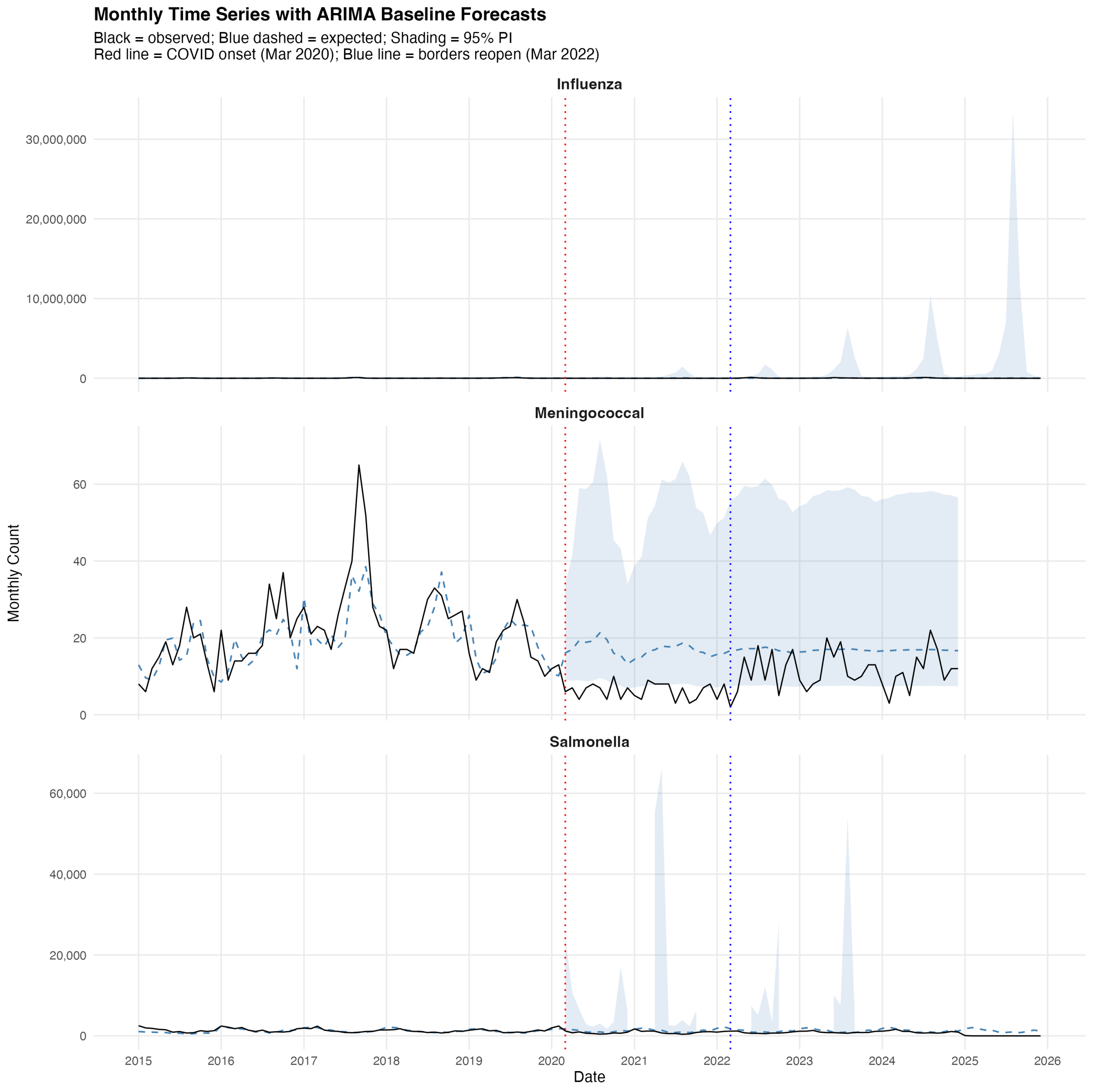


**Figure S3:** Figure S3. Monthly time series with ARIMA baseline forecasts

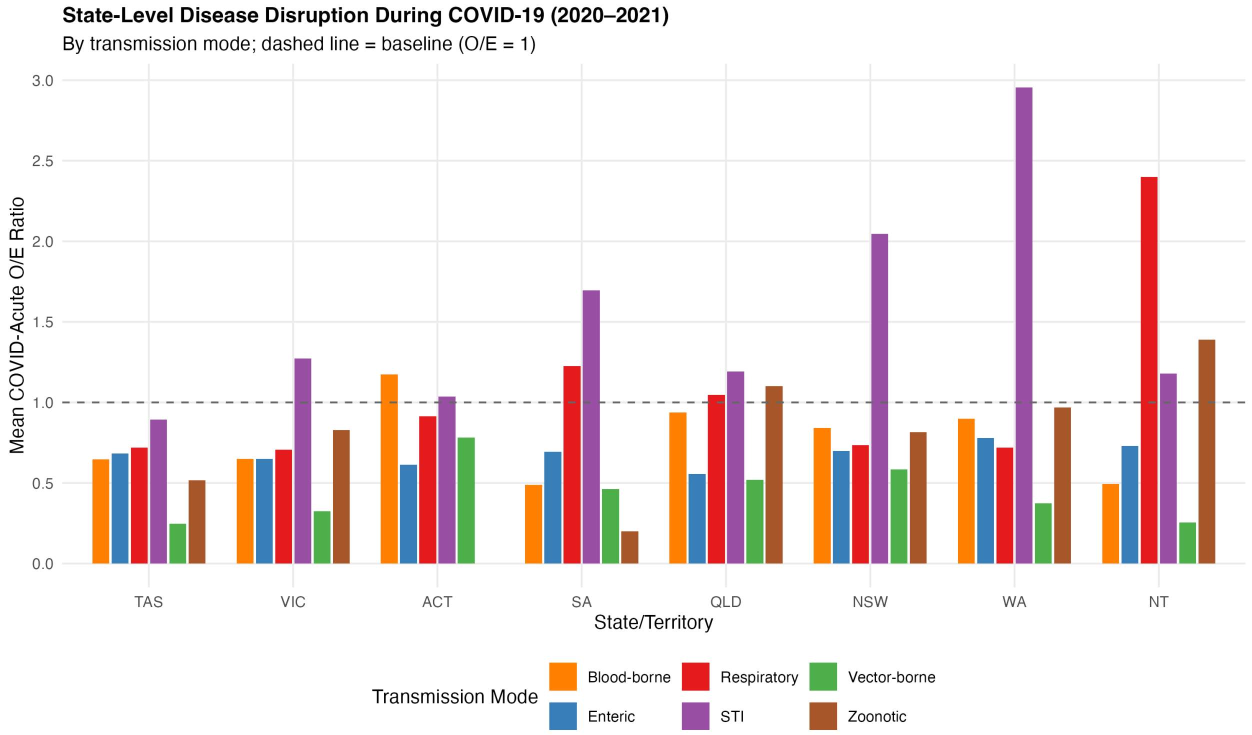


**Figure S4:** Figure S4. State-level comparison of disruption

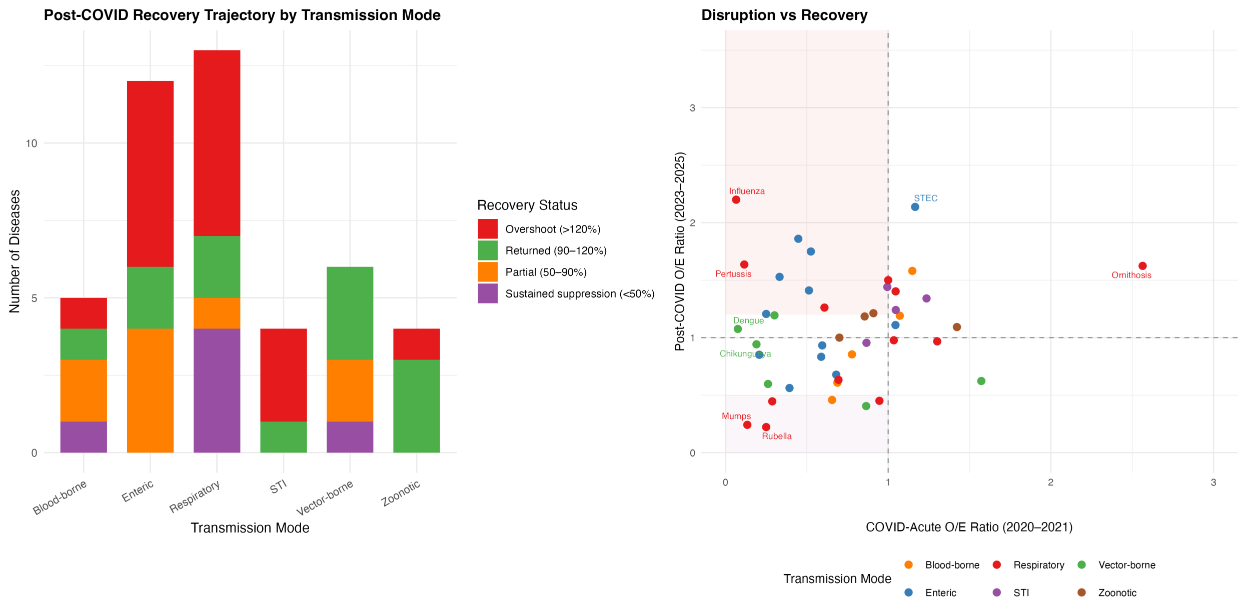


**Figure S5:** Figure S5. Recovery trajectories

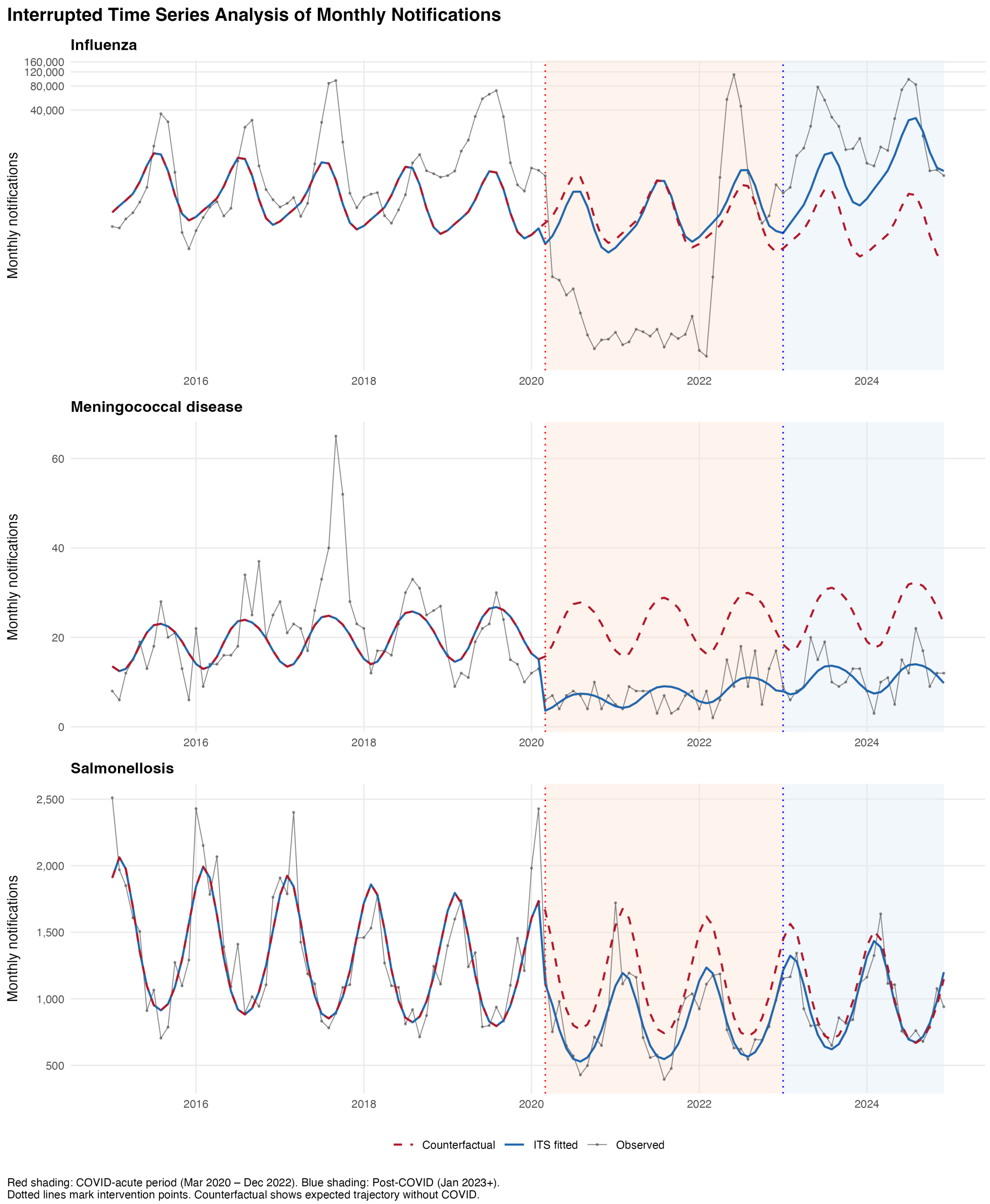


**Figure S6:** Figure S6. Monthly interrupted time series

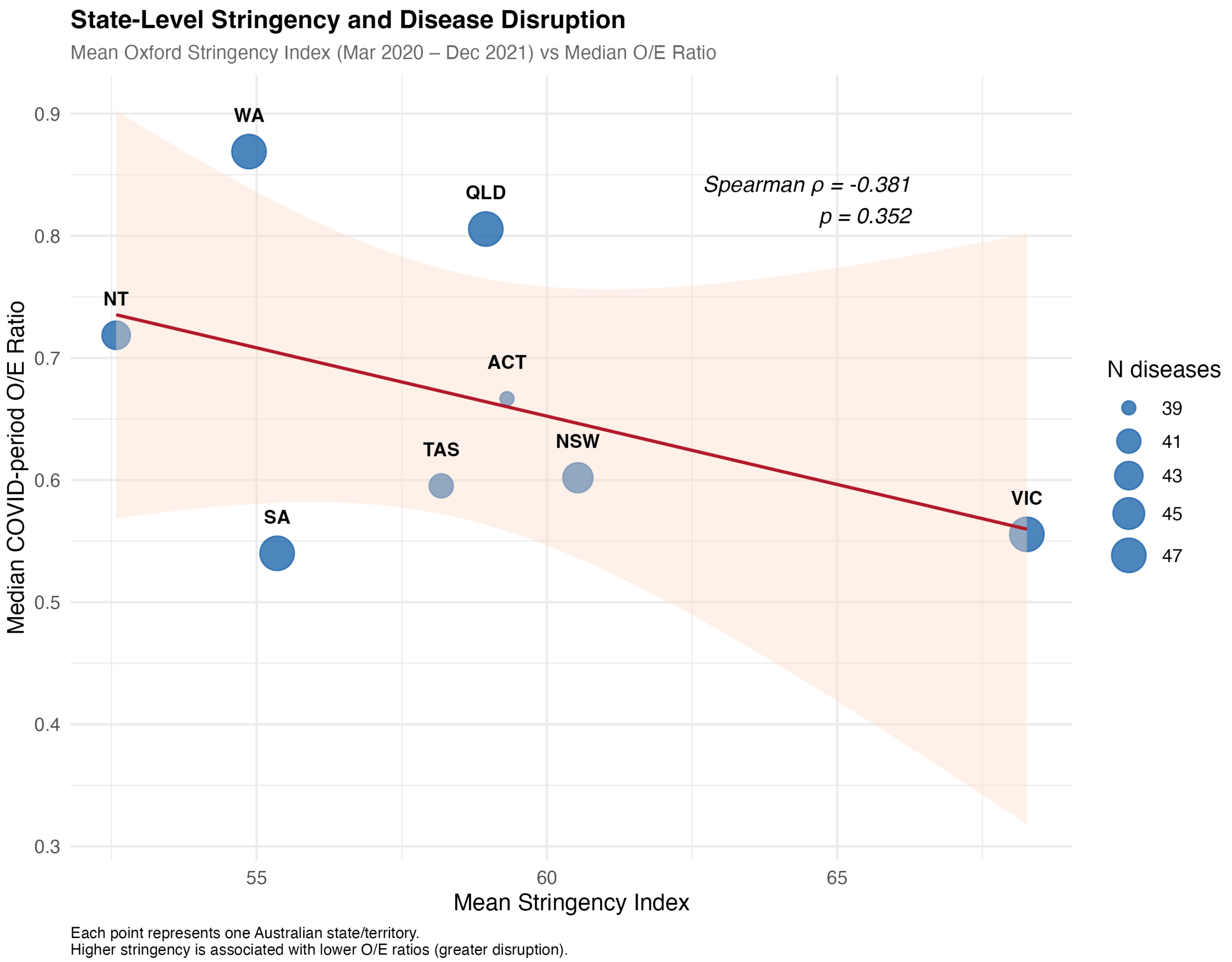


**Figure S7:** Figure S7. State-level NPI stringency scatter plot

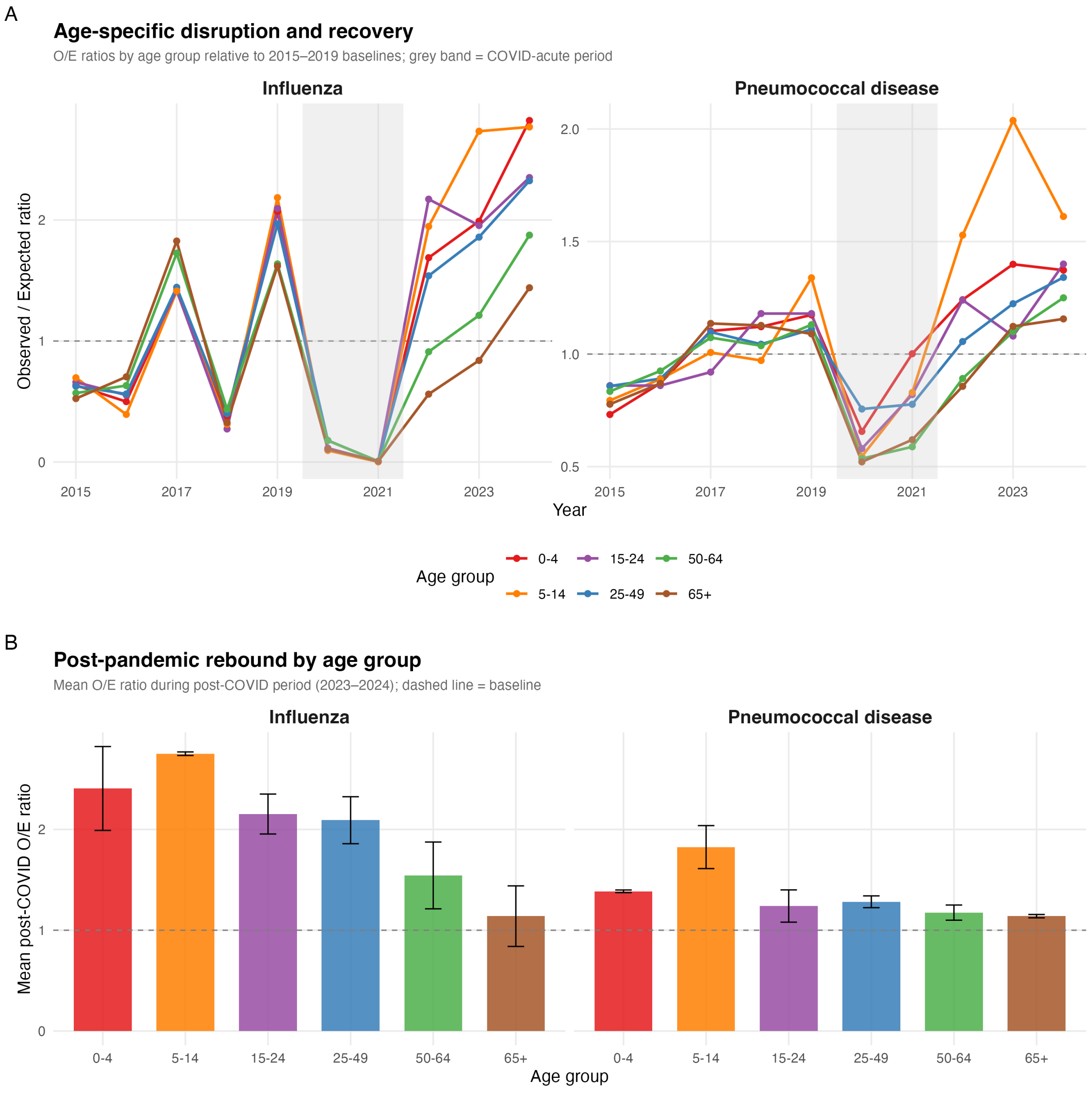


**Figure S8:** Figure S8. Age-specific disruption and recovery

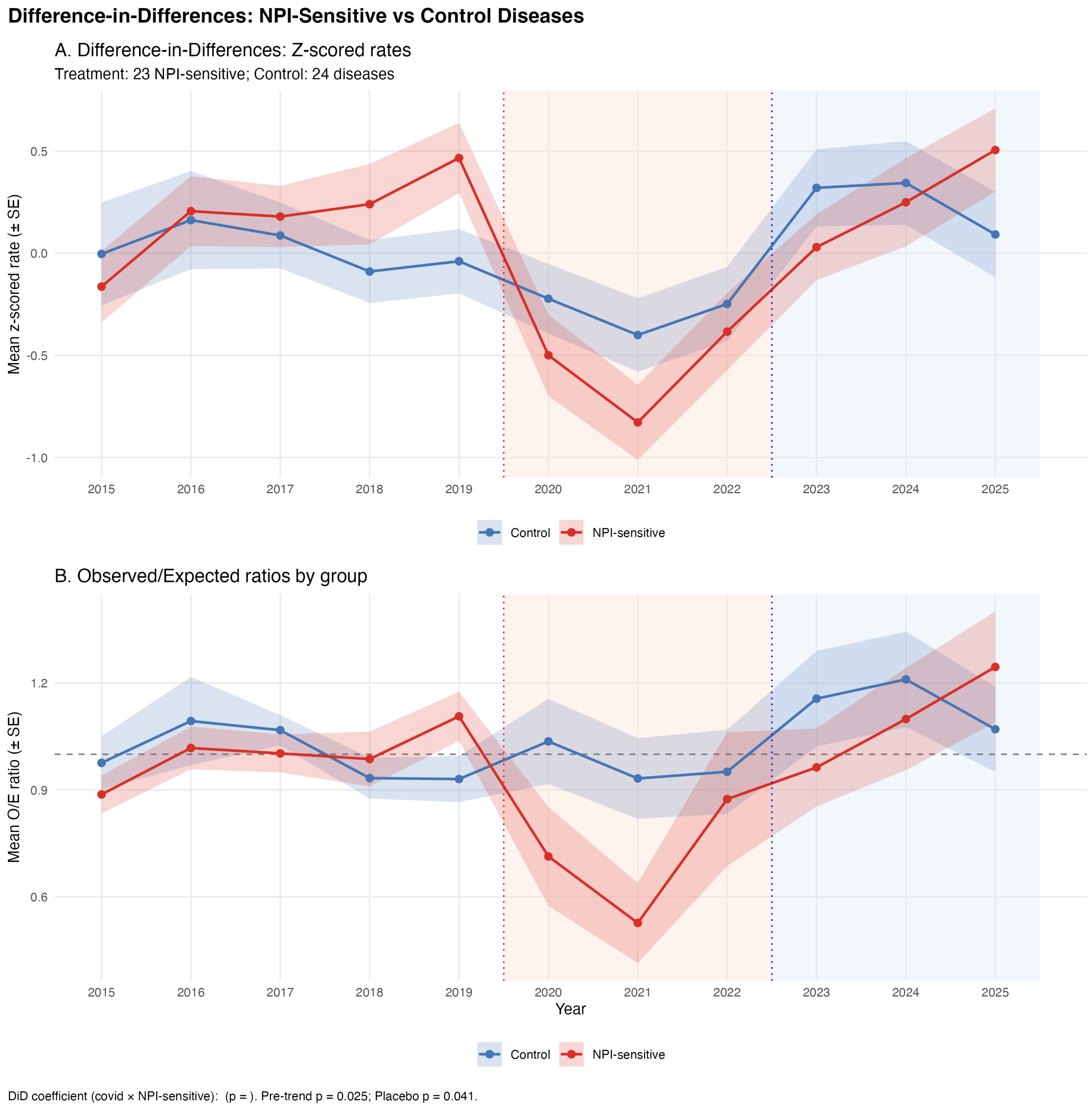


**Figure S9:** Figure S9. DiD analysis — NPI-sensitive specification (secondary)

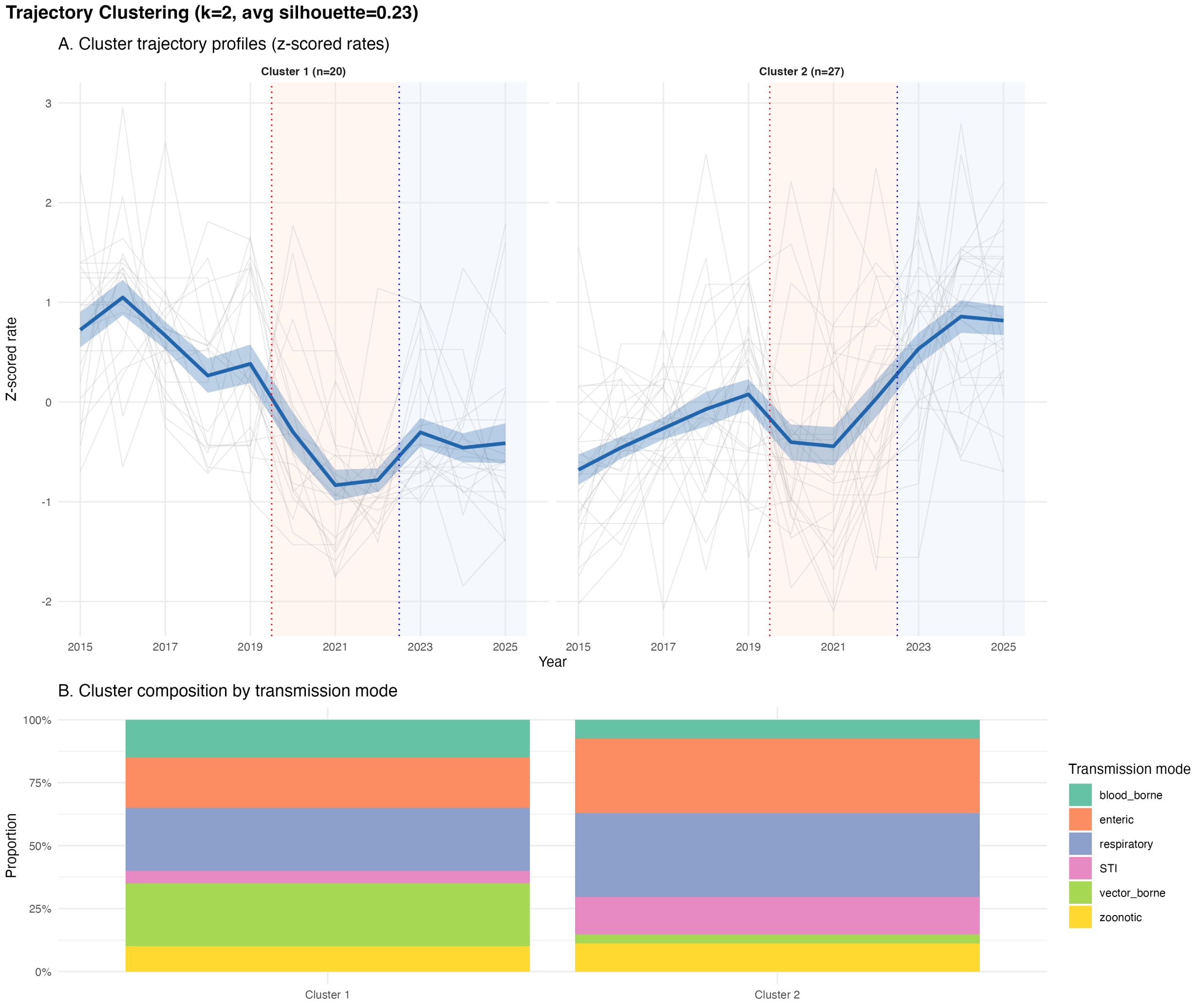


**Figure S10:** Figure S10. Disruption severity group profiles (k-medoids, k=2)

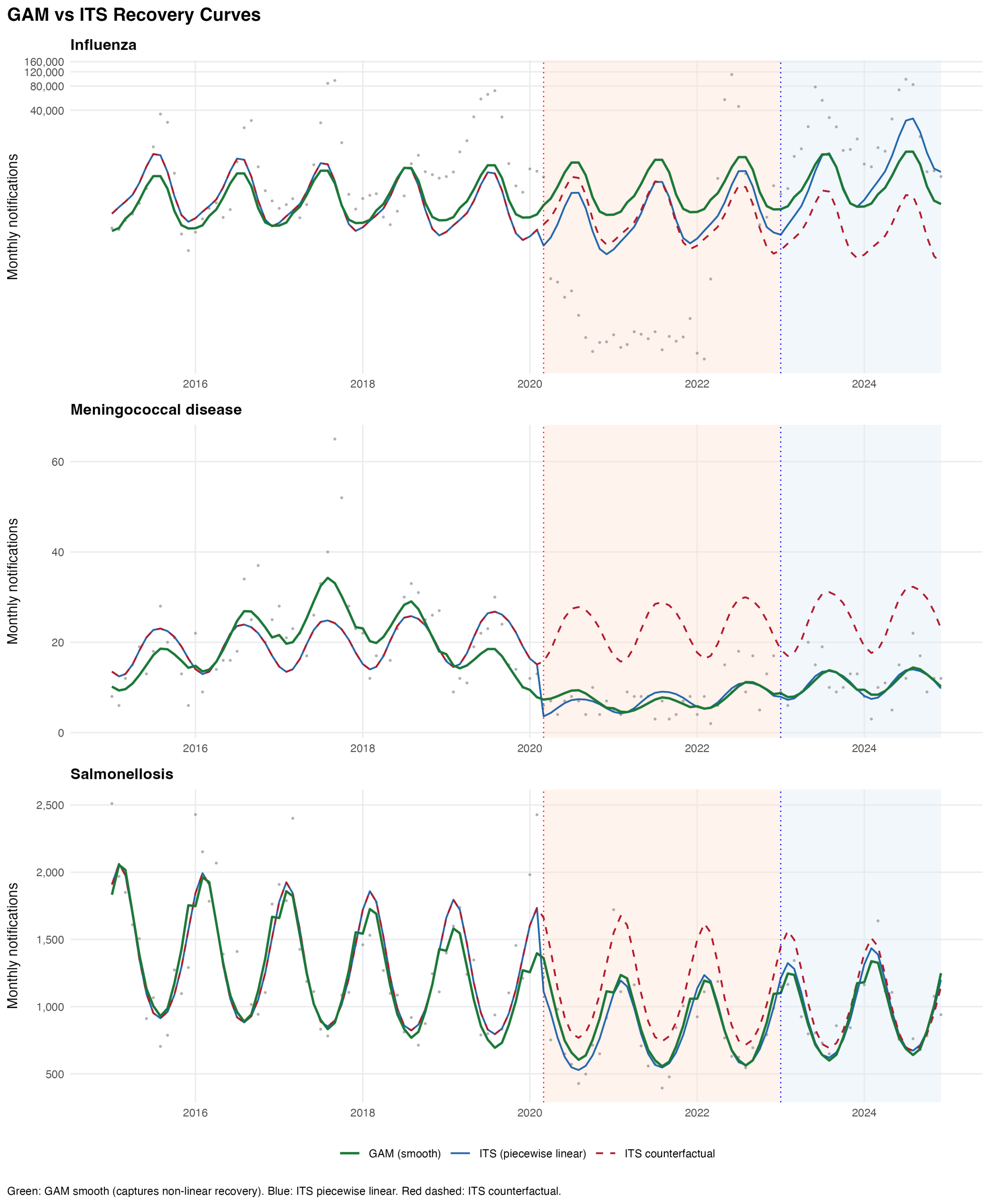


**Figure S11:** Figure S11. GAM recovery curves
